# Associations of severe COVID-19 with polypharmacy in the REACT-SCOT case-control study

**DOI:** 10.1101/2020.07.23.20160747

**Authors:** Paul M McKeigue, Sharon Kennedy, Amanda Weir, Jen Bishop, Stuart J McGurnaghan, David McAllister, Chris Robertson, Rachael Wood, Nazir Lone, Janet Murray, Thomas M Caparrotta, Alison Smith-Palmer, David Goldberg, Jim McMenamin, Colin Ramsay, Bruce Guthrie, Sharon Hutchinson, Helen M Colhoun On behalf of Public Health Scotland COVID-19 Health Protection Study Group.

## Abstract

**Objectives:** To investigate the relation of severe COVID-19 to prior drug prescribing.

**Design:** Matched case-control study (REACT-SCOT) based on record linkage to hospital discharges since June 2015 and dispensed prescriptions issued in primary care during the last 240 days.

**Setting:** Scottish population.

**Main outcome measure:** Severe COVID-19, defined by entry to critical care or fatal outcome.

**Participants:** All 4272 cases of severe COVID-19 in Scotland since the start of the epidemic, with 36948 controls matched for age, sex and primary care practice.

**Results:** Severe COVID-19 was strongly associated with the number of non-cardiovascular drug classes dispensed. This association was strongest in those not resident in care homes, in whom the rate ratio (95% CI) associated with dispensing of 12 or more drug classes versus none was 10.8 (8.7, 13.2), and was not accounted for by treatment of conditions designated as conferring increased risk. Of 17 drug classes postulated at the start of the epidemic to be “medications compromising COVID”, all were associated with increased risk of severe COVID-19. The largest effect was for antipsychotic agents: rate ratio 4.14 (3.39, 5.07). Other drug classes with large effects included proton pump inhibitors (rate rato 2.19 (1.70, 2.80) for >= 2 defined daily doses/day), opioids (3.62 (2.65, 4.94) for >= 50 mg morphine equivalent/day) and gabapentinoids. These associations persisted after adjusting for covariates, and were stronger with recent than with non-recent exposure.

**Conclusions:** Severe COVID-19 is associated with polypharmacy and with drugs that cause sedation, respiratory depression or dyskinesia, have anticholinergic effects or affect the gastrointestinal system. These associations are not easily explained by co-morbidity. Although the evidence for causality is not conclusive, these results support existing guidance on reducing overprescribing of these drug classes and limiting inappropriate polypharmacy as a potential means of reducing COVID-19 risk.

**Registration:** ENCEPP number EUPAS35558

**What is already known on this topic:** Two previous studies have examined the relationship of severe COVID-19 to drugs for the cardiovascular system. This is the first systematic study of the relationship of severe COVID-19 to prior drug prescribing.

**What this study adds:** Severe COVID-19 is associated with polypharmacy and with drugs that cause sedation, respiratory depression or dyskinesia, have anticholinergic effects or affect the gastrointestinal system. These associations are not easily explained by co-morbidity. These results support earlier warnings that these drug classes that these drugs might increase susceptibility to COVID-19, and reinforce existing guidance on reducing overprescribing of these drug classes.

## Background

In the initial analysis of REACT-SCOT, a matched case control study of risk factors for severe COVID-19 in Scotland, we reported a strong association of severe COVID-19 with having had at least one prescription dispensed in the past year^1^. The univariate rate ratios associated with at least one prescription varied from 3.8 in those aged under 60 years to 2.3 in those aged 75 years and over. This association persisted after adjusting for care home residence, hospital admission in the last five years, and diagnoses of conditions designated by public health agencies as conferring vulnerability to COVID-19 (hereafter listed conditions). The objective of this study was to investigate this association.

## Methods

The design of the REACT-SCOT case-control study has been described in detail elsewhere^1^. As this study was set up in response to a national emergency no arrangements were made for patient and public involvement. All individuals testing positive for nucleic acid for SARS-CoV-2 in Scotland were ascertained through the Electronic Communication of Surveillance in Scotland (ECOSS) database. Admissions to critical care were obtained from the Scottish Intensive Care Society and Audit Group (SICSAG) database that captures admission to all critical care (intensive care or high dependency) units. Death registrations were obtained from linkage to the National Register of Scotland. Severe or fatal COVID-19 was defined by either (1) a positive nucleic acid test followed by entry to critical care or death within 28 days; or (2) a death certificate with COVID-19 as underlying cause.

For each case, the Community Health Index database was used to select up to ten controls matched for sex, one-year age band and registered with the same primary care practice, who were alive on the same day as the first date that the case tested positive. For fatal cases who had not tested positive, the incident date was assigned as 14 days before death. For this analysis based on ascertainment of positive test results up to 6 June 2020, entry to critical care up to 14 June 2020 and deaths up to 12 June 2020 there were 4272 cases and 36948 controls.

### Morbidity and drug prescribing

For all cases and controls, ICD-10 diagnostic codes were extracted from the last five years of hospital discharge records in the Scottish Morbidity Record (SMR01), excluding records of discharges less than 25 days before testing positive for SARS-CoV-2, and from the national cancer registry. Diagnoses of diabetes were extracted from linkage to the national diabetes register. British National Formulary (BNF) drug codes for dispensed prescriptions issued in primary care were extracted from the Scottish Prescribing Information System^2^. A cutoff date of 15 days before the incident date (date of testing positive for SARS-CoV-2, or 14 days before death for fatal cases without a positive test) was set, and prescriptions dispensed in a 240-day interval before this cutoff date were included. For this analysis prescription codes from BNF chapters 14 and above, comprising dressings, appliances, vaccines, anaesthesia and other preparations were grouped as “Other”.

We began by testing for association of severe COVID-19 with the number of drug classes (BNF subparagraph codes) for which at least one prescription had been dispensed during the period of observation. In accordance with earlier suggestions that prescribing of multiple cardiovascular drugs, which is supported by evidence-based guidelines, should be considered separately from putatively inappropriate polypharmacy^3^, we partitioned the number of drug classes dispensed into cardiovascular and other drugs. To define a prespecified hypothesis about drug classes postulated to increase the risk of severe COVID-19, we used a review by Laporte and Healy published on 2 April 2020 that listed 17 widely prescribed drug classes associated with increased pneumonia risk and therefore of concern as “medications compromising COVID”^4^. Excluding immunosuppressive drugs that are criteria for shielding, the Laporte-Healy list comprised proton pump inhibitors, gastrointestinal antispasmodics, H1 antihistamines, hypnotics and sedatives, antipsychotic drugs, antidepressants, drugs used in nausea and vertigo, opioid analgesics, gabapentinoids, anti-epileptic drugs, antimuscarinic drugs used in parkinsonism, urinary antispasmodics, and non-steroidal anti-inflammatory drugs.

For proton pump inhibitors and opioids, it was possible to use equivalent doses to calculate a total dose for all drugs in the class. Defined daily doses (DDDs) of each proton pump inhibitor were obtained from the DDD/ATC Index of the WHO Collaborating Centre for Drug Statistics Methodology. Morphine milligram equivalents (MMEs) for opioids were obtained from the website of the Faculty of Pain Medicine of the Royal College of Anaesthestists. Average daily doses of proton pump inhibitors (as DDDs) and opioids (as MME) were calculated for each individual as the sum of (conversion factor *×* strength *×* quantity dispensed) divided by observation period. The dose of opioid was calculated as the sum over opioid-containing items in subparagraph 0407010 (nonopioid and compound preparations) and 0407020 (opioid analgesics).

As described previously, we derived indicator variables for a list of conditions that have been designated as risk conditions for COVID-19 by public health agencies^5^: diabetes, heart disease, asthma or chronic obstructive airway disease, chronic kidney disease, disabling neurological disease, liver disease and immunodeficiency or immunosuppression. ICD-10 diagnostic and BNF drug codes used to derive these conditions are available with the [ENCEPP registration](http://www.encepp.eu/encepp/openAttachment/documents.otherDocument-0/35565). Socioeconomic status was encoded as the quintile of the postcode-based Scottish Index of Multiple Deprivation (SIMD).

## Statistical analysis

Rate ratios for severe COVID-19 (hereafter COVID-19) were estimated from conditional logistic regression models, implemented as Cox regression in the R function `survival::clogit`. To distinguish between causality and confounding, we used several approaches:

- Testing for consistency of association with drugs that have a similar mode of action across different indication groups
- Testing for a dose-response relationship and stratifying by age group
- Adjusting for prespecified covariates.
- Comparison between recent (last 120 days of observation) and less recent (1 June 2019 to 121 days before exposure) time windows of dispensing.

## Results

### Relation of severe COVID-19 to polypharmacy

Figure 1 shows that the rate of of severe COVID-19 increased steeply with the number of non-cardiovascular drug classes and decreased with the number of noncardiovascular drug classes dispensed. Table 1 shows that this association was restricted to those not resident in care homes. Supplementary Table S1 shows that among those not resident in care homes, the association of severed COVID-19 with number of non-cardiovascular drug classes dispensed was present both in those without any diagnosed listed condition and in those with at least one diagnosed listed condition.

**Fig 1.**
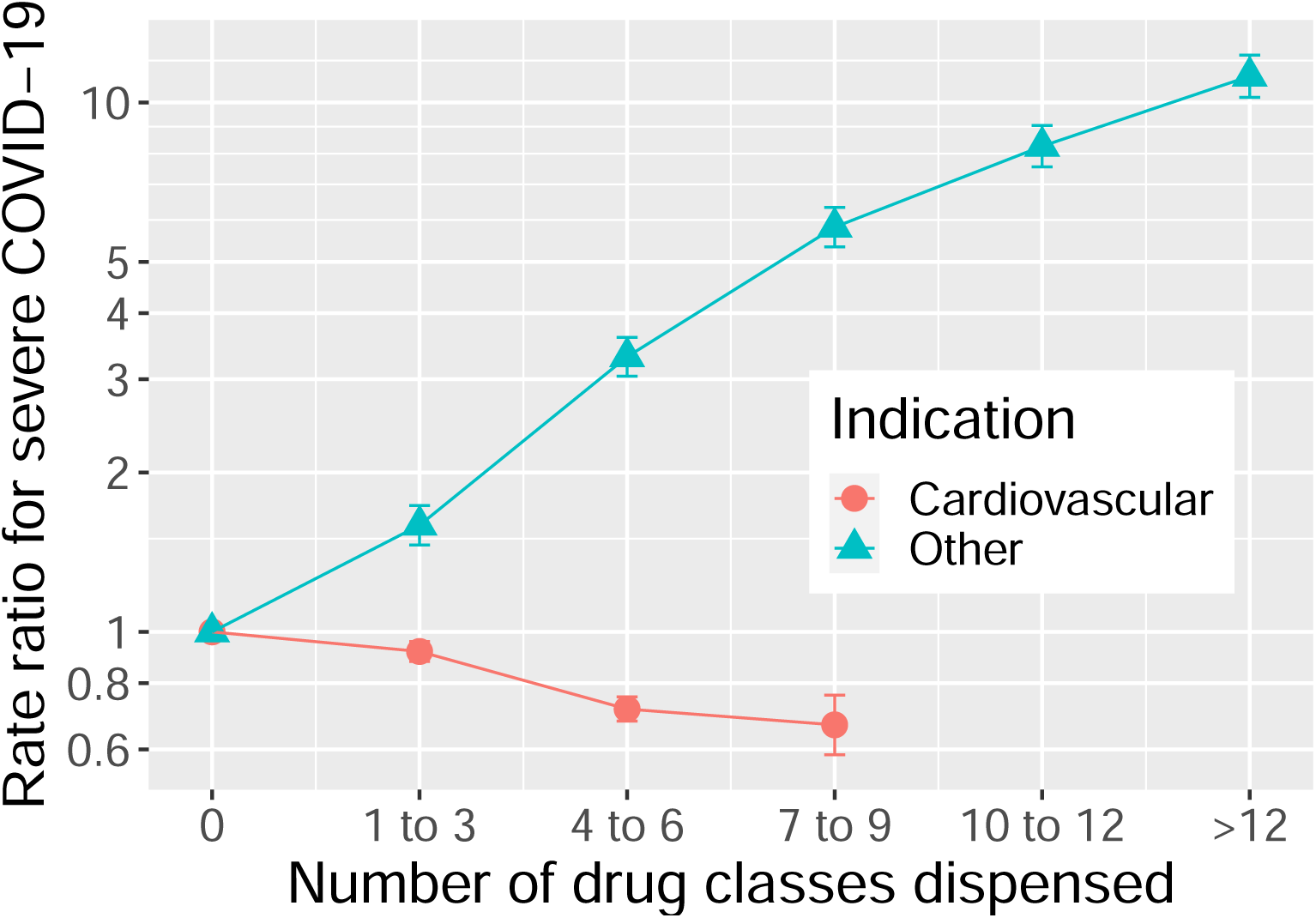
Rate ratios (with standard errors) in a conditional logistic regression of severe COVID-19 on number of cardiovascular (BNF chapter 4) and non-cardiovascular drug classes dispensed

**Table 1.**
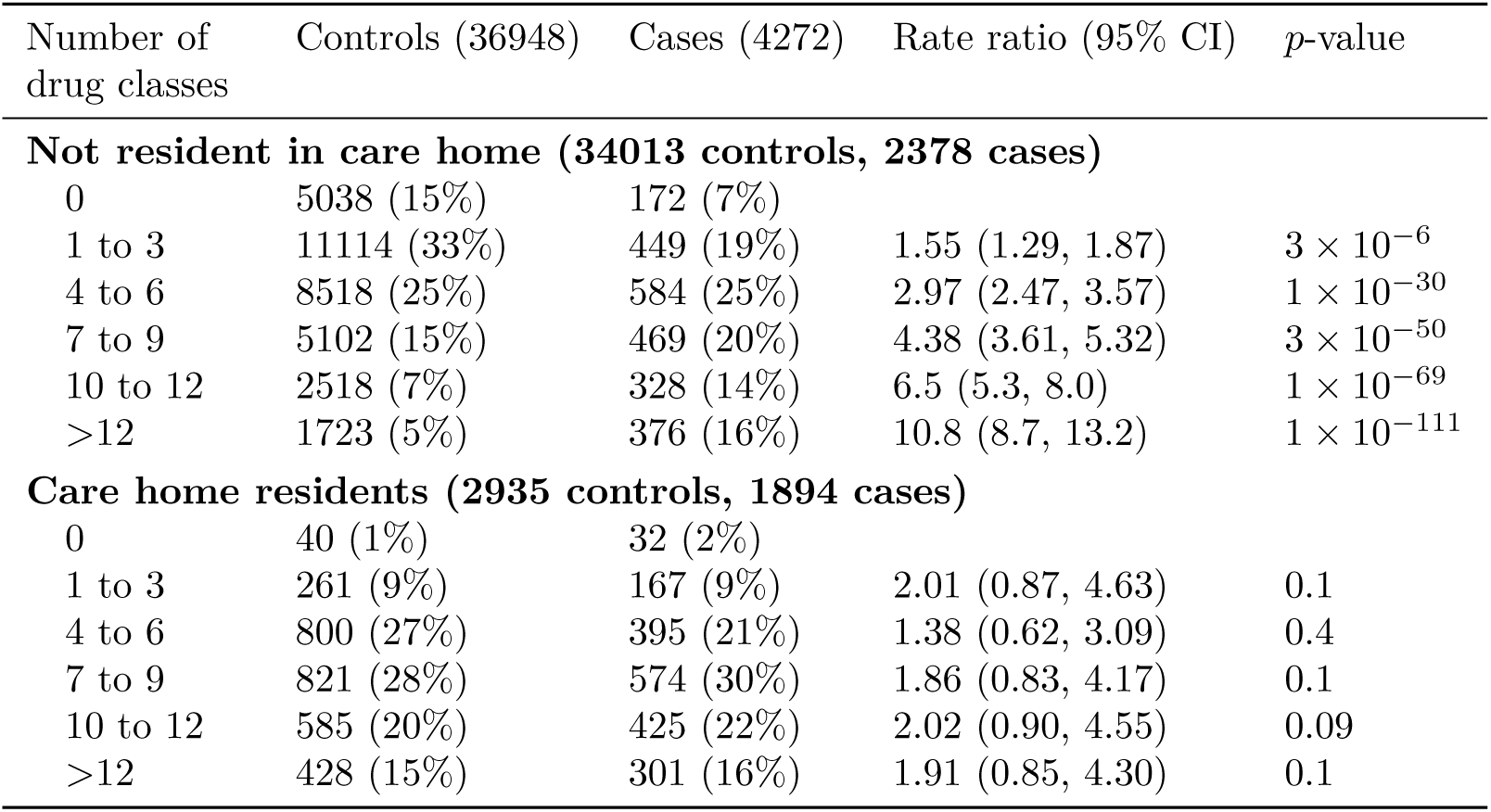
Association of severe COVID-19 with number of non-cardiovascular drug classes dispensed, by care home residence

| Number of drug classes | Controls (36948) | Cases (4272) | Rate ratio (95% CI) | *p*-value |
| --- | --- | --- | --- | --- |
| **Not resident in care home (34013 controls, 2378 cases)** |  |  |  |  |
| 0 | 5038 (15%) | 172 (7%) |  |  |
| 1 to 3 | 11114 (33%) | 449 (19%) | 1.55 (1.29, 1.87) | $3 \times 10^{-6}$ |
| 4 to 6 | 8518 (25%) | 584 (25%) | 2.97 (2.47, 3.57) | $1 \times 10^{-30}$ |
| 7 to 9 | 5102 (15%) | 469 (20%) | 4.38 (3.61, 5.32) | $3 \times 10^{-50}$ |
| 10 to 12 | 2518 (7%) | 328 (14%) | 6.5 (5.3, 8.0) | $1 \times 10^{-69}$ |
| >12 | 1723 (5%) | 376 (16%) | 10.8 (8.7, 13.2) | $1 \times 10^{-111}$ |
| **Care home residents (2935 controls, 1894 cases)** |  |  |  |  |
| 0 | 40 (1%) | 32 (2%) |  |  |
| 1 to 3 | 261 (9%) | 167 (9%) | 2.01 (0.87, 4.63) | 0.1 |
| 4 to 6 | 800 (27%) | 395 (21%) | 1.38 (0.62, 3.09) | 0.4 |
| 7 to 9 | 821 (28%) | 574 (30%) | 1.86 (0.83, 4.17) | 0.1 |
| 10 to 12 | 585 (20%) | 425 (22%) | 2.02 (0.90, 4.55) | 0.09 |
| >12 | 428 (15%) | 301 (16%) | 1.91 (0.85, 4.30) | 0.1 |

### Associations with specific drug classes

In order to identify specific drug classes associated with severe COVID-19, we examined thse associations among those not resident in a care home and without any listed condition, as most people diagnosed with listed conditions such as asthma and diabetes are on medication for these conditions. Table 2 shows univariate associations of dispensing of at least one drug in each BNF subparagraph, filtered to show only drug classes with at least 50 exposed individuals and *p <* 0.001. The drug classes associated with severe COVID-19 include proton pump inhibitors, laxatives, multiple classes of drugs acting on the central nervous system, nutritional supplements, and non-steroidal anti-inflammatory drugs.

**Table 2.** Univariate rate ratios for severe COVID-19 associated with dispensing of each BNF subparagraph code, restricted to those without a listed condition and not resident in a care home, filtered to show only drug classes with at least 50 exposed individuals and *p <* 0.001:

| BNF subparagraph | Controls<br>(17280) | Cases<br>(650) | Rate ratio | $p$-value |
| --- | --- | --- | --- | --- |
| 103050: Proton pump inhibitors | 4670 | 232 | 1.84 (1.52, 2.22) | $4 \times 10^{-10}$ |
| 104020: Antimotility drugs | 264 | 20 | 2.72 (1.54, 4.81) | $6 \times 10^{-4}$ |
| 105010: Aminosalicylates | 149 | 15 | 3.42 (1.78, 6.60) | $2 \times 10^{-4}$ |
| 106040: Osmotic laxatives | 1255 | 78 | 2.28 (1.67, 3.11) | $2 \times 10^{-7}$ |
| 401020: Anxiolytics | 488 | 41 | 2.12 (1.43, 3.16) | $2 \times 10^{-4}$ |
| 402010: Antipsychotic drugs | 166 | 25 | 3.89 (2.29, 6.62) | $5 \times 10^{-7}$ |
| 403010: Tricyclic and related<br>antidepressant drugs | 1042 | 64 | 1.85 (1.35, 2.52) | $1 \times 10^{-4}$ |
| 403040: Other antidepressant drugs | 503 | 48 | 2.68 (1.85, 3.88) | $2 \times 10^{-7}$ |
| 407010: Non-opioid analgesics and<br>compound preparations | 4221 | 224 | 2.01 (1.65, 2.45) | $4 \times 10^{-12}$ |
| 407020: Opioid analgesics | 1032 | 80 | 2.32 (1.73, 3.13) | $3 \times 10^{-8}$ |
| 408010: Control of epilepsy | 750 | 51 | 2.26 (1.59, 3.20) | $5 \times 10^{-6}$ |
| 501012: Penicillinase-resistant<br>penicillins | 574 | 41 | 2.26 (1.52, 3.36) | $6 \times 10^{-5}$ |
| 501013: Broad-spectrum penicillins | 1160 | 71 | 1.99 (1.47, 2.69) | $1 \times 10^{-5}$ |
| 603020: Use of corticosteroids | 333 | 33 | 3.36 (2.11, 5.35) | $3 \times 10^{-7}$ |
| 901011: Oral iron | 409 | 26 | 2.81 (1.66, 4.78) | $1 \times 10^{-4}$ |
| 901020: Drugs used in megaloblastic<br>anaemias | 1023 | 56 | 2.10 (1.48, 2.97) | $3 \times 10^{-5}$ |
| 904020: Enteral nutrition | 99 | 17 | 5.8 (2.7, 12.2) | $5 \times 10^{-6}$ |
| 906040: Vitamin D | 1732 | 73 | 1.83 (1.32, 2.55) | $3 \times 10^{-4}$ |
| 1001010: Non-steroidal<br>anti-inflammatory drugs | 1454 | 86 | 1.65 (1.27, 2.14) | $2 \times 10^{-4}$ |

Table 3 shows associations of severe COVID-19 with dispensing of each of the drug classes on the Laporte-Healy list in those not resident in a care home. All drug classes on this list were associated with increased risk in univariate analyses (though the association with NSAIDS was stronger in Table 2 where listed conditions were excluded. This included all the drug classes listed by Laporte and Healy as having anticholinergic effects: H1 antihistamines, antidepressants, urinary antispasmodics, gastrointestinal antispasmodics, drugs for vertigo, antimuscarinic drugs used in the treatment of parkinsonism, and antiepileptic drugs.

**Table 3.**
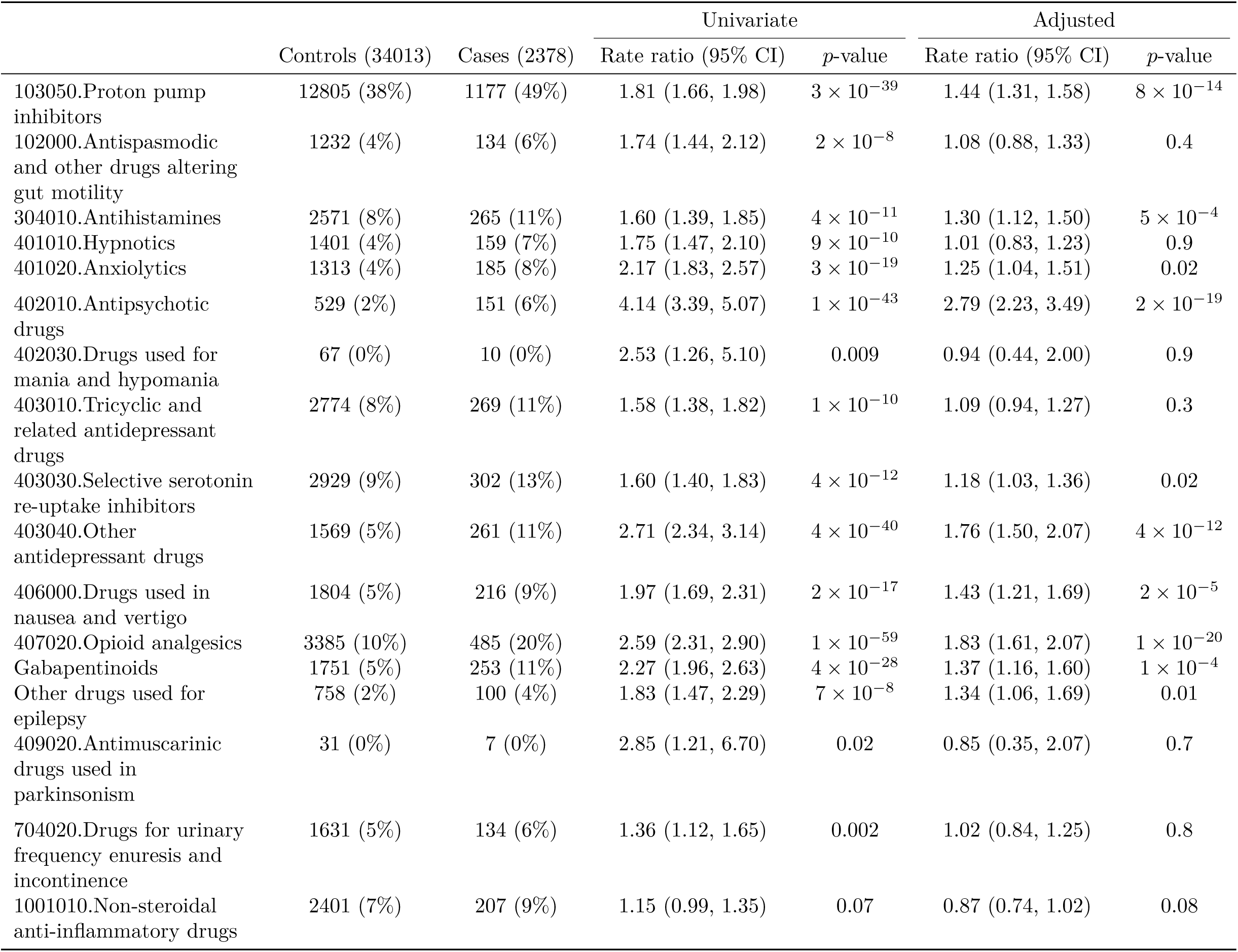
Associations of severe COVID-19 with drug classes listed by Laporte and Healy (2020), restricted to those not resident in a care home. Adjusted rate ratios are based on a joint model with all drug classes in the table and SIMD quintile as covariates

In a multivariable regression, the strongest independent associations were with proton pump inhibitors, antihistamines, antipsychotic drugs, and opioid analgesics. In both univariate and multivariable analyses, the highest rate ratio was that associated with antipsychotic drugs. As others have noted^4^, the chemical structures and modes of action of drugs used in the treatment of nausea and vertigo overlap with those of antipsychotic drugs. Supplementary Table S2 shows the univariate associations of severe COVID-19 with specific drugs classified in these two subparagraphs of the BNF. Across both groups of indications, phenothiazines and other drugs that are dopamine antagonists were strongly associated with increased rates of severe disease. Rate ratios were elevated both for phenothiazines and for second-generation antipsychotics: aripiprazole, olanzapine, risperidone and amrisulpride.

### Time window analyses

Supplementary Table S3 shows for each drug class on the Laporte-Healy list the rate ratio associated with dispensing in the most recent 120-day time window, with dispensing only in the previous time window as reference category. Because most users of these drugs had dispensed prescriptions in both time windows, these analyses are based on relatively small numbers: the drug classes shown are restricted to those with at least 500 cases and controls exposed only in the earlier time window. For most of these drug classes shown, the rate ratio associated with recent exposure only is above 1, but only for opioid analgesics, and to some extent proton pump inhibitors and anxiolytics, does this association reach conventional levels of statistical significance.

### Dose-response analyses

Supplementary Table S4 shows the relationship of severe COVID-19 to average daily doses of opioids (as MME) and proton pump inhibitors (as DDDs) over the 240-day observation period. For both these drug classes there were dose-response relationships, and for proton pump inhibitors this relationship was strongest in those aged less than 75 years. With unexposed as baseline, the univariate rate ratio associated with opioid use in this age group was 3.62 (2.65, 4.94) in those with average daily dose of more than 50 mg morphine equivalent (MME), reduced to 2.90 (2.04, 4.11) on adjusting for care home residence, SIMD quintile and any history of neoplasm. For proton pump inhibitors, the univariate rate ratio associated with average dose of 2 or more DDDs / day was 2.19 (1.70, 2.80), reduced to 1.98 (1.50, 2.60) by adjusting for care home residence, SIMD quintile, any diagnosis of ICD-10 codes K20-K31 (diseases of esophagus, stomach and duodenum), non-steroidal anti-inflammatory drugs, anti-platelet agents and anticoagulants.

### Associations with other drug classes

Supplementary Tables S5 and S6 show associations with drug classes in BNF chapters 2 (cardiovascular) and 10 (musculoskeletal). These chapters were selected as of interest because because specific hypotheses about possible effects of drugs in these chapters – ACE inhibitors^6^, anticoagulants^7^, and hydroxychloroquine^8^– have been proposed or discussed.

Table S5 shows associations with drugs for the cardiovascular system. Prescriptions of loop diuretics and anticoagulants were associated with elevated rate ratios for severe COVID-19 in univariate and multivariate analyses. Over all age groups combined, the univariate rate ratio associated with oral anticoagulants was reduced from 1.88 (1.66, 2.12) to 1.28 (1.12, 1.46) by adjustment for ischaemic heart disese, other heart disease and ever-use of a proton pump inhibitor. Of drug classes commonly used to treat hypertension, ACE inhibitors and angiotensin II receptor antagonists were associated with reduced risk of COVID-19.

Table S6 shows associations with drugs for the musculoskeletal system disaggregated by generic name, as the BNF groups all disease-modifying antirheumatic drugs under a single subparagraph code. All antirheumatic drugs were associated with elevated rate ratios for COVID-19. The univariate rate ratio associated with hydroxychloroquine sulfate was 2.10 (1.34, 3.28); adjustment for ever-use of a proton pump inhibitor reduced this to 1.80 (1.15, 2.82).

## Discussion

We have shown that severe COVID-19 is associated with polypharmacy, defined by the number of drugs classes dispensed during the period of observation, in individuals without conditions designated as conferring high risk. The rate ratios of 5 to 7 associated with dispensing of more than 10 drug classes are larger than the ratio of about 2 for all-cause mortality associated with this level of polypharmacy in a systematic review^9^. Attempting to investigate associations with specific drugs with a hypothesis-free approach is difficult because many of the drug classes that are strongly associated with severe COVID-19, such as proton pump inhibitors, opioids and gabapentinoids, are indicators of overprescribing, recognized as such in the Scottish National Therapeutic Index of prescribing quality^10^.

To narrow the hypothesis space we tested a pre-specified list of drugs postulated at the start of the epidemic to increase risk of severe COVID-19, based on previously described associations with pneumonia or activity on relevant pathways, especially anticholinergic agents^4^. We have shown that all the drugs originally listed are associated with increased risk. Distinguishing between causality and confounding as possible explanations for the association of COVID-19 with these drug classes is difficult because drug exposures are not measured accurately, and the associations are likely to be confounded by other drugs, by co-morbidities and more generally by frailty and socioeconomic deprivation.

The strongest association was with antipsychotic drugs. As the recognized indications for these drugs are not known to be associated with susceptibility to severe COVID-19, there is no obvious explanation for this association other than causality. Further evidence favouring causality is that drugs prescribed for nausea that have similar chemical structures and mode of action to antipsychotic drugs show similar associations with severe COVID-19.

For both opioids and proton pump inhibitors, four criteria provide moderate but not conclusive evidence favouring causal explanations over confounding. There are strong dose-response relationships of COVID-19 to dispensed average daily dose. Adjustment for covariates pre-specified as likely to confound these associations reduces the effect size only slightly. The effect sizes are larger in younger individuals and when the analysis is restricted to those without any of the designated risk conditions for COVID-19. Among ever-exposed individuals, the rate ratios associated with dispensing only in the most recent time window were higher than the rate ratios associated with dispensing only in an earlier time window. The dose-response relationship of opioid use to COVID-19 is similar in magnitude to that reported for community-acquired pneumonia in a study of people receiving medical care through the Veterans Administration from 2000-2012^11^. Although gabapentinoids are classified in the BNF under subparagraph 0408010 (“Control of epilepsy”), in Scotland they are widely used in combination with or as substitutes for opioid analgesics.

A limitation of this study is that we do not have morbidity data from primary care, which would include risk factors such as smoking and coding of presenting complaints. Another limitation is that it is not possible to capture hospital prescribing data which includes biologic agents that have immunosuppressive effects. Strengths of this study are that diagnoses are based on hospital discharge records coded to ICD-10 (rather than the SNOMED-CT codes used in primary care databases), and that drug exposure is based on dispensed rather than issued prescriptions.

The mechanisms postulated by Laporte and Healy for drugs to increase risk of severe COVID-19 include sedation, respiratory depression, respiratory dyskinesia and anticholinergic effects^4^. We note that as SARS-CoV-2 is at least partly an enteric infection^12^ and the ACE2 receptor is expressed in the intestine, it is plausible that proton pump inhibitors and other drugs acting on the gastrointestinal tract could increase susceptibility to severe infection. It may be relevant to investigate these associations in other countries where COVID-19 epidemics have been especially severe and overprescribing of drug classes such as proton pump inhibitors^13–15^ or opioids^16^ has been reported previously.

We emphasize that because of the relationship of COVID-19 to polypharmacy, associations with specific drug classes cannot be studied without taking into account how those drug classes are related to the profile of drug prescribing. Of those drug classes that are not on the Laporte-Healy list, there are strong univariate associations of severe COVID-10 with dispensing of antibiotics, laxatives and nutritional supplements. The associations with nutritional supplements is likely to be confounded by overprescribing. For ACE inhibitors and angiotensin-II receptor blockers, our results are consistent with other studies that have found no increased risk associated with these drugs^6,17^ and indeed suggest some protective effect may be possible. To explore this more fully will require access to other datasets where measurements of blood pressure and other covariates are available. The relation of anticoagulant use to COVID-19 is of interest because coagulopathy is a feature of severe disease^7^. Although in this study anticoagulants were associated with increased risk in univariate analysis, this association was reduced by adjusting for covariates including diagnosed heart disease and co-prescribing of proton pump inhibitors. Associations with disease-modifying antirheumatic drugs such as hydroxychloroquine are likely to be confounded by hospital-based prescribing of biologic anti-rheumatic drugs, which is not captured by record linkage in Scotland.

### Conclusion

Severe COVID-19 is strongly associated with polypharmacy. This association is not easily explained by co-morbidity, and it is strongest in those without hospital diagnoses of conditions that confer high risk of disease. As a prediction of which drug classes would be associated with increased susceptibility to severe COVID-19, the Laporte-Healy list prepared at the start of the epidemic appears to be remarkably accurate. Many of the drug classes on this list are recognized indicators of overprescribing. The consistency of associations with drugs that have similar modes of action across different groups of indication, the dose-response and time window effects support causal explanations for at least some these associations. We recommend that public health agencies should reinforce existing guidelines on avoidance of overprescribing of these drug classes and more generally on inappropriate polypharmacy.

## Declarations

### Ethics approval and information governance

This study was conducted under approvals from the Public Benefit and Privacy Panel for Health and Social Care that allow Public Health Scotland staff to link datasets. Datasets were de-identified before analysis.

### Data sharing

Requests for acccess to the data may be submitted to the Public Benefit and Privacy Panel.

### Transparency declaration

PM, as the manuscript’s guarantor, affirms: that the manuscript is an honest, accurate, and transparent account of the study being reported; that no important aspects of the study have been omitted; and that any discrepancies from the study as originally planned and registered have been explained. This manuscript has been generated directly from the source data by a reproducible research pipeline. All source code used for derivation of variables, statistical analysis and generation of this manuscript is freely available on [GitHub](http://github.com/pmckeigue/covid-scotland_public).

### Registration

The study protocol was registered with the European Network of Centres for Pharmacoepidemiology and Pharmacovigilance (ENCEPP number [EUPAS35558](http://www.encepp.eu/encepp/viewResource.htm?id=35559)).

## Data Availability

Applications for access to the data may be made to the Public Benefit and Privacy Panel for Health and Social Care, Scotland

## Acknowledgements

We thank all staff in critical care units who submitted data to the SICSAG database, the Scottish Morbidity Record Data Team, the staff of the National Register of Scotland, the Public Health Scotland Terminology Services, the HPS COVID-19 Laboratory & Testing cell and the NHS Scotland Diagnostic Virology Laboratories, and Nicola Rowan (HPS) for coordinating this collaboration.

## Public Health Scotland COVID-19 Health Protection Study Group

Alice Whettlock^1^, Allan McLeod^1^, Andrew Gasiorowski^1^, Andrew Merrick^1^, Andy McAuley^1^, April Went^1^, Calum Purdie^1^, Colin Fischbacher^1^, Colin Ramsay^1^, David Bailey^1^, David Henderson^1^, Diogo Marques^1^, Eisin McDonald^1^, Genna Drennan^1^, Graeme Gowans^1^, Graeme Reid^1^, Heather Murdoch^1^, Jade Carruthers^1^, Janet Fleming^1^, Jade Carruthers^1^, Joseph Jasperse^1^, Josie Murray^1^, Karen Heatlie^1^, Lindsay Mathie^1^, Lorraine Donaldson^1^, Martin Paton^1^, Martin Reid^1^, Melissa Llano^1^, Michelle Murphy-Hall^1^, Paul Smith^1^, Ros Hall^1^, Ross Cameron^1^, Susan Brownlie^1^, Adam Gaffney^2^, Aynsley Milne^2^, Christopher Sullivan^2^, Edward McArdle^2^, Elaine Glass^2^, Johanna Young^2^, William Malcolm^2^, Jodie McCoubrey ^2^

1 Health Protection Scotland (Public Health Scotland), Meridian Court, 5 Cadogan Street, Glasgow G2 6QE.

2 NHS National Services Scotland, Meridian Court, 5 Cadogan Street, Glasgow G2 6QE.

### Supplementary material

The R and Rmarkdown scripts used to generate this article, which include the code used to derive variables, will be made available when the paper is published.

## Supplementary Tables

**Table S1.** Association of severe COVID-19 with number of non-cardiovascular drug classes dispensed, in those not resident in a care home by diagnosis of at least one listed condition

| Number of drug classes | Controls (34013) | Cases (2378) | Rate ratio (95% CI) | *p*-value |
| --- | --- | --- | --- | --- |
| **No listed condition (17280 controls, 650 cases)** |  |  |  |  |
| 0 | 4534 (26%) | 125 (19%) |  |  |
| 1 to 3 | 7149 (41%) | 229 (35%) | 1.59 (1.25, 2.02) | $1 \times 10^{-4}$ |
| 4 to 6 | 3565 (21%) | 153 (24%) | 2.69 (2.04, 3.55) | $2 \times 10^{-12}$ |
| 7 to 9 | 1425 (8%) | 79 (12%) | 4.11 (2.88, 5.88) | $9 \times 10^{-15}$ |
| 10 to 12 | 458 (3%) | 46 (7%) | 7.3 (4.6, 11.7) | $3 \times 10^{-17}$ |
| >12 | 149 (1%) | 18 (3%) | 7.2 (3.8, 13.8) | $3 \times 10^{-9}$ |
| **At least one listed condition (16733 controls, 1728 cases)** |  |  |  |  |
| 0 | 504 (3%) | 47 (3%) |  |  |
| 1 to 3 | 3965 (24%) | 220 (13%) | 0.62 (0.43, 0.89) | 0.01 |
| 4 to 6 | 4953 (30%) | 431 (25%) | 1.01 (0.71, 1.43) | 1 |
| 7 to 9 | 3677 (22%) | 390 (23%) | 1.33 (0.93, 1.90) | 0.1 |
| 10 to 12 | 2060 (12%) | 282 (16%) | 1.87 (1.29, 2.69) | $9 \times 10^{-4}$ |
| >12 | 1574 (9%) | 358 (21%) | 2.95 (2.05, 4.25) | $5 \times 10^{-9}$ |

**Table S2.** Univariate associations of severe COVID-19 with drugs in four subparagraphs of BNF chapter 4, in those not resident in a care home and not treated for cancer in last year, filtered to retain rows with at least 20 exposed individuals

|  | Controls (31024) | Cases (2081) | Rate ratio (95% CI) | *p*-value |
| --- | --- | --- | --- | --- |
| **Antipsychotic drugs** |  |  |  |  |
| Aripiprazole | 38 (0%) | 12 (1%) | 3.19 (1.61, 6.33) | $9 \times 10^{-4}$ |
| Olanzapine | 96 (0%) | 18 (1%) | 2.63 (1.55, 4.49) | $4 \times 10^{-4}$ |
| Levomepromazine | 27 (0%) | 17 (1%) | 9.9 (4.7, 20.6) | $1 \times 10^{-9}$ |
| Risperidone | 123 (0%) | 40 (2%) | 4.90 (3.26, 7.38) | $3 \times 10^{-14}$ |
| Haloperidol | 18 (0%) | 9 (0%) | 8.8 (3.3, 23.0) | $1 \times 10^{-5}$ |
| Chlorpromazine hydrochloride | 26 (0%) | 7 (0%) | 3.00 (1.28, 7.03) | 0.01 |
| Amisulpride | 21 (0%) | 4 (0%) | 6.0 (1.7, 21.4) | 0.005 |
| **Drugs for nausea and vertigo** |  |  |  |  |
| Prochlorperazine | 770 (2%) | 59 (3%) | 1.36 (1.02, 1.81) | 0.03 |
| Cinnarizine | 150 (0%) | 12 (1%) | 1.46 (0.78, 2.71) | 0.2 |
| Cyclizine | 283 (1%) | 44 (2%) | 2.74 (1.93, 3.88) | $2 \times 10^{-8}$ |
| Metoclopramide hydrochloride | 159 (1%) | 24 (1%) | 2.60 (1.62, 4.16) | $7 \times 10^{-5}$ |
| Betahistine dihydrochloride | 368 (1%) | 17 (1%) | 0.68 (0.41, 1.13) | 0.1 |
| Domperidone | 54 (0%) | 10 (0%) | 2.63 (1.27, 5.43) | 0.009 |
| **Opioids** |  |  |  |  |
| Morphine | 454 (1%) | 121 (6%) | 4.53 (3.58, 5.73) | $2 \times 10^{-36}$ |
| Codeine phosphate | 567 (2%) | 82 (4%) | 2.65 (2.03, 3.46) | $8 \times 10^{-13}$ |
| Tramadol hydrochloride | 1144 (4%) | 119 (6%) | 1.73 (1.41, 2.13) | $2 \times 10^{-7}$ |
| Fentanyl | 102 (0%) | 14 (1%) | 2.17 (1.18, 4.00) | 0.01 |
| Buprenorphine | 154 (0%) | 9 (0%) | 1.18 (0.57, 2.45) | 0.6 |
| Oxycodone | 249 (1%) | 55 (3%) | 4.27 (3.05, 5.99) | $3 \times 10^{-17}$ |
| **Gabapentinoids** |  |  |  |  |
| Gabapentin | 974 (3%) | 115 (6%) | 1.90 (1.54, 2.35) | $2 \times 10^{-9}$ |
| Lamotrigine | 143 (0%) | 21 (1%) | 2.22 (1.37, 3.60) | 0.001 |
| **Control of epilepsy** |  |  |  |  |
| Topiramate | 34 (0%) | 5 (0%) | 1.96 (0.74, 5.17) | 0.2 |
| Carbamazepine | 176 (1%) | 14 (1%) | 1.17 (0.67, 2.06) | 0.6 |
| Clonazepam | 46 (0%) | 10 (0%) | 2.97 (1.44, 6.11) | 0.003 |
| Levetiracetam | 137 (0%) | 24 (1%) | 2.62 (1.64, 4.17) | $5 \times 10^{-5}$ |
| Sodium valproate | 106 (0%) | 9 (0%) | 1.00 (0.50, 2.01) | 1 |
| Phenobarbital | 20 (0%) | 4 (0%) | 2.28 (0.70, 7.39) | 0.2 |
| Phenytoin | 49 (0%) | 11 (1%) | 3.08 (1.54, 6.17) | 0.001 |
| Primidone | 29 (0%) | 4 (0%) | 1.91 (0.64, 5.70) | 0.2 |
| Lacosamide | 19 (0%) | 2 (0%) | 1.52 (0.34, 6.79) | 0.6 |

**Table S3.**
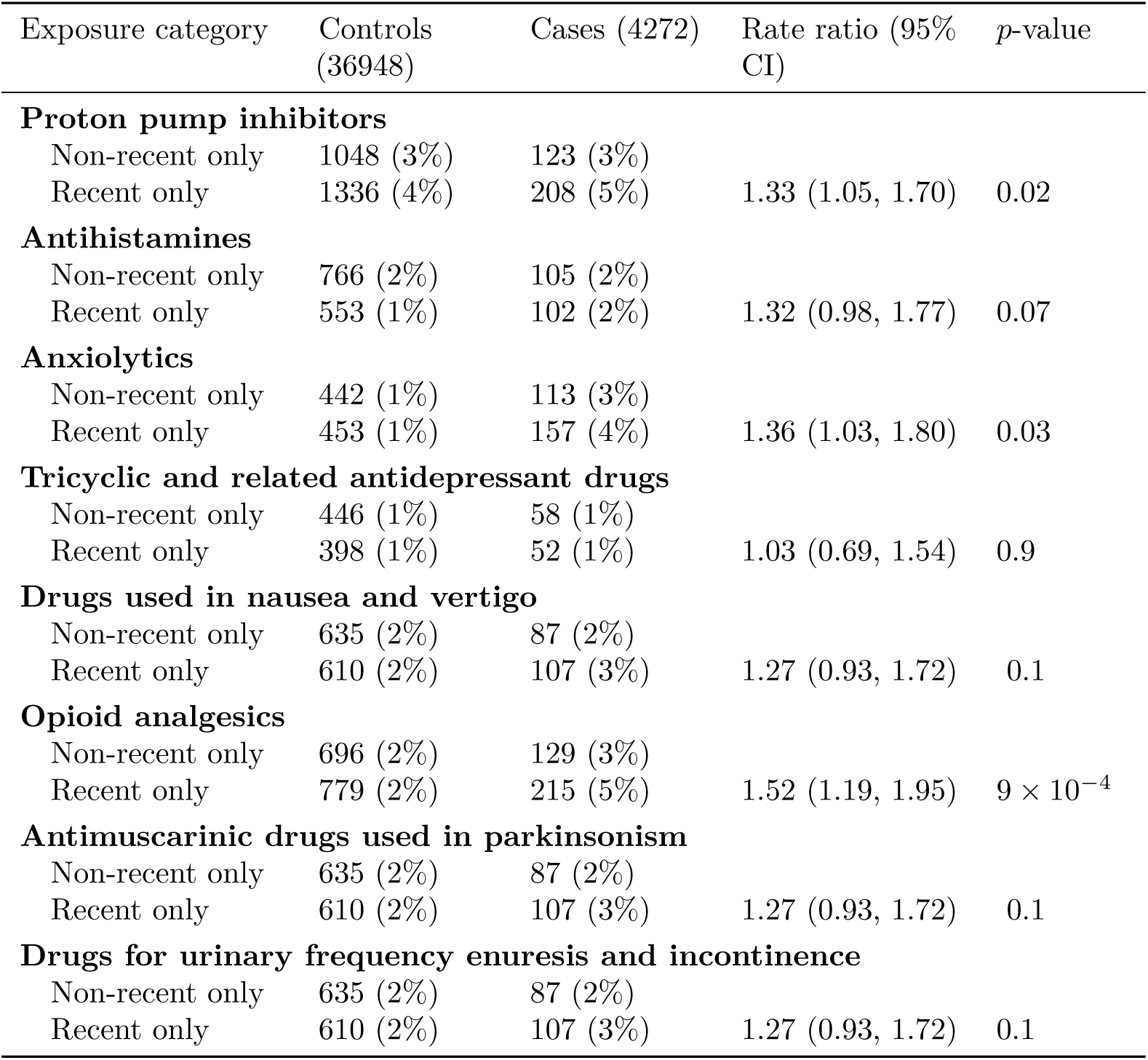
Comparison of associations with exposure only in the most recent 120-day time window with exposure only in the previous 120-day time window (baseline category for rate ratios) for drug classes on the Laporte-Healy list with at least 500 cases and controls exposed only in the earlier time window

**Table S4.**
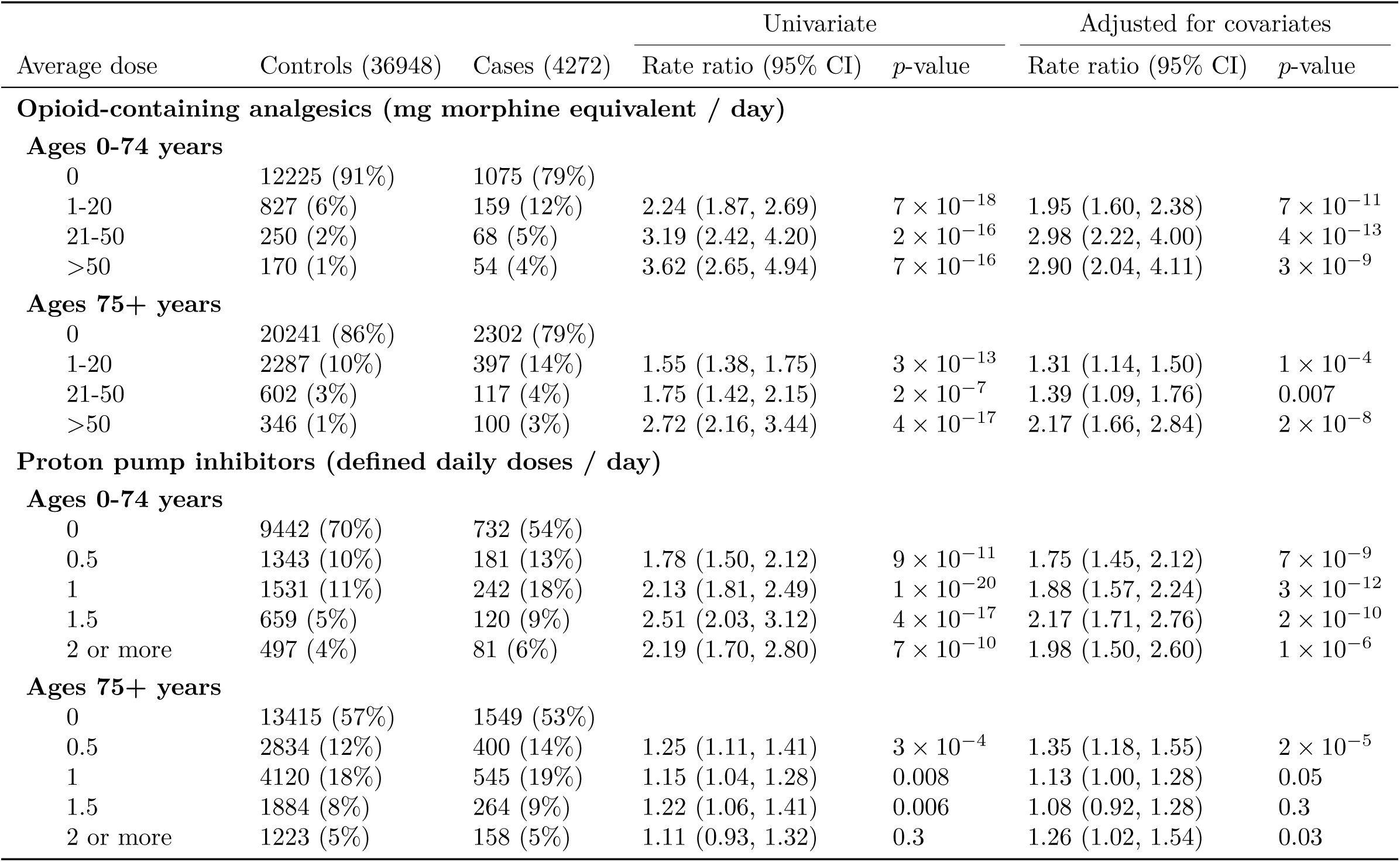
Associations of severe COVID-19 with dispensed average daily dose of opioids and proton pump inhibitors, with and without adjustment for prespecified covariates

**Table S5.**
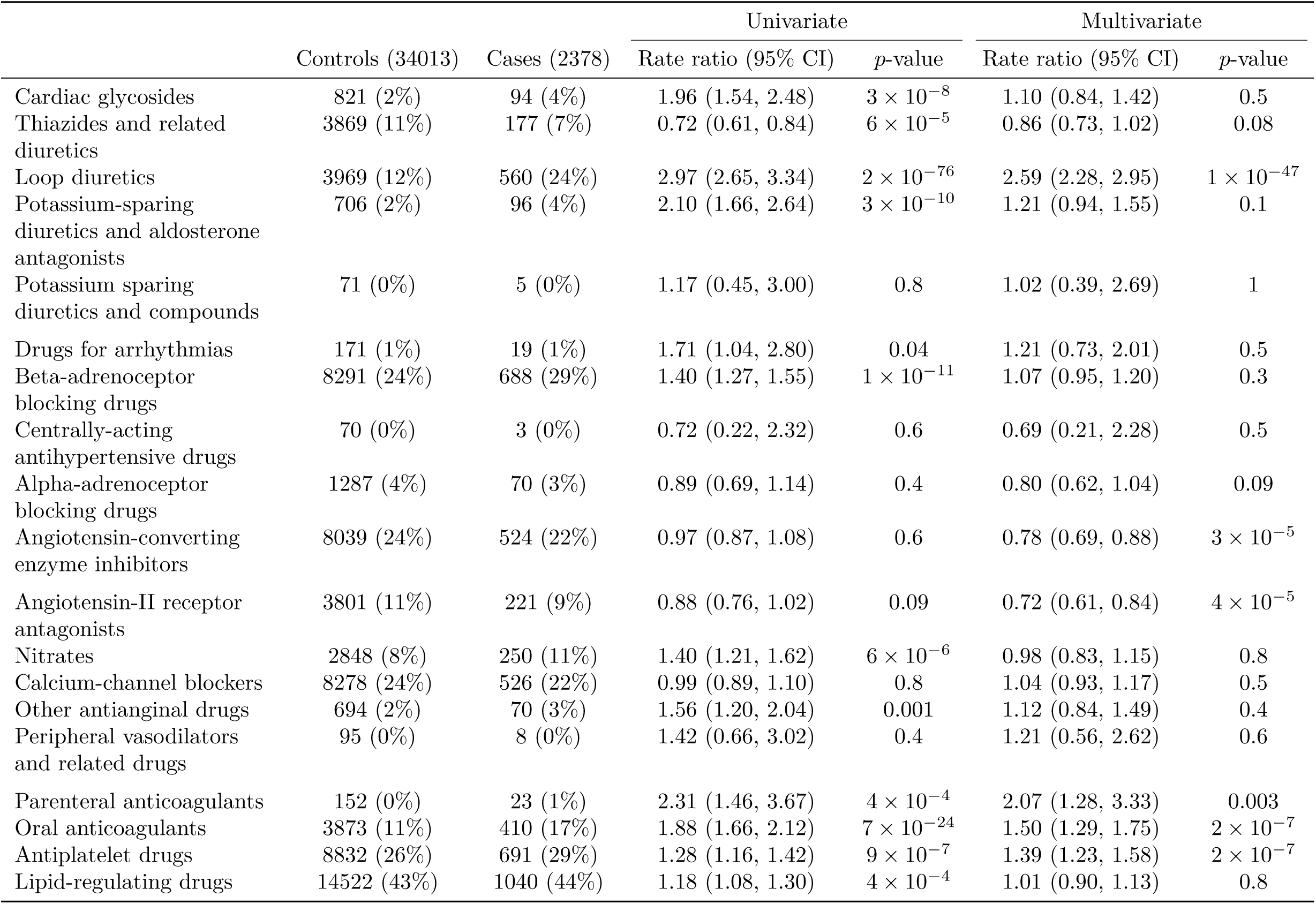
Associations of severe COVID-19 with prescribed drugs in BNF chapter 2 by subparagraph code, restricted to those not resident in a care home and filtered to retain rows with at least 50 exposed individuals

**Table S6.**
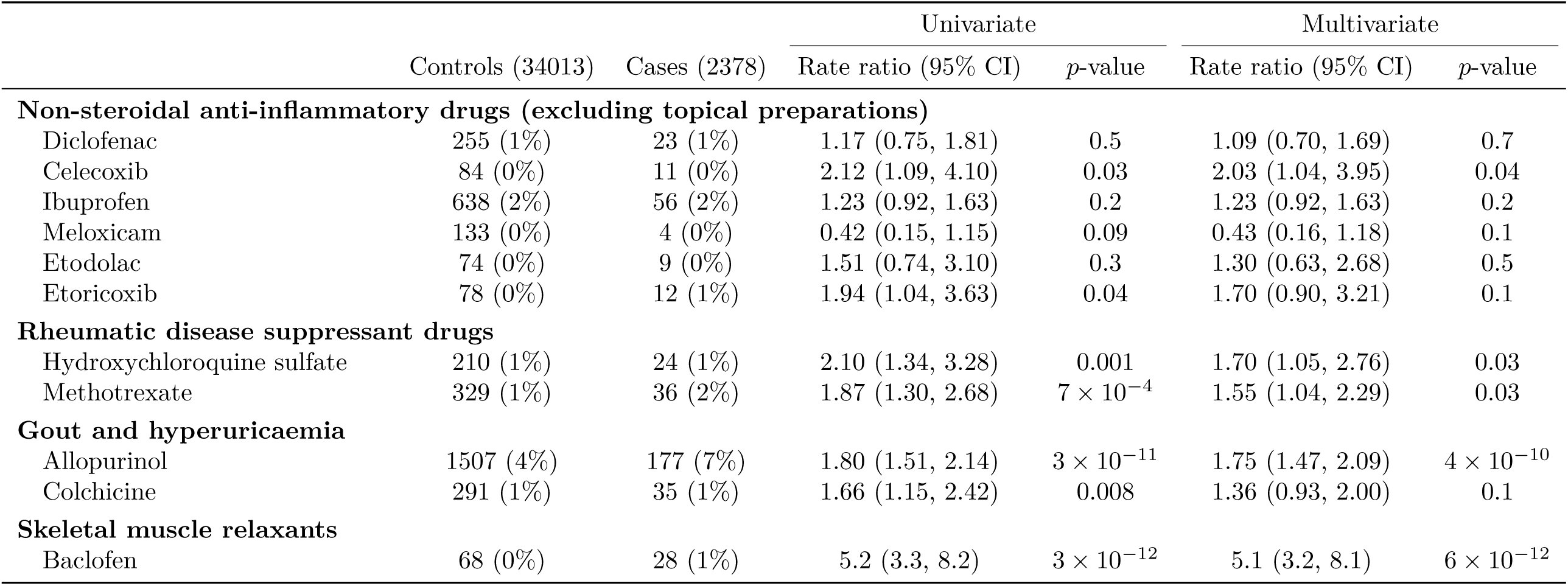
Associations of severe COVID-19 in with prescribed drugs in BNF chapter 10 by approved name, restricted to those not resident in a care home and filtered to retain rows with at least 50 exposed individuals

